# Estimation of Transmission Potential and Severity of COVID–19 in Romania and Pakistan

**DOI:** 10.1101/2020.05.02.20088989

**Authors:** Muhammad Ozair, Takasar Hussain, Mureed Hussain, Aziz Ullah Awan, Dumitru Baleanu

## Abstract

During the outbreak of an epidemic, it is of immense interest to monitor the effects of containment measures and forecast of outbreak including epidemic peak. To confront the epidemic, a simple *SIR* model is used to simulate the number of affected patients of Coronavirus disease in Romania and Pakistan. The model captures the growth in case onsets and the estimated results are almost compatible with the actual reported cases. Through the calibration of parameters, forecast for the appearance of new cases in Romania and Pakistan is reported till the end of this year by analysing the current situation. The constant level of number of patients and time-to-reach this level is also reported through the simulations. The drastic condition is also discussed which may occur if all the preventive restraints are removed.

## 1 Introduction

In December (2019), the Wuhan Municipal Health Commission (Hubei Province, China) informed to World Health Organization (WHO) about a group of 27 cases of unknown etiology pneumonia, who were commonly exposed to a fish and live animals market in Wuhan City. It was also notified that seven of these patients were critically serious. The symptoms of the first case began on December 8, 2019. On January 7, 2020, Chinese authorities identified a new type of family virus as the agent causing the outbreak. The causative agent of this pneumonia was identified as a new virus in the *Coronaviridae* family that has since been named SARS–CoV–2. The clinical picture associated with this virus has been named *COVID* – 19. On march 11, WHO declared the global pandemic [1]. The worldwide reported cases of *COVID* – 19 are ~ 3 million with nearly 0.2 million deaths.

Coronaviruses are a family of viruses that cause infection in humans and some animals. Diseases by coronavirus are zoonotic, that is, they can be transmitted from animals to humans [2]. Coronaviruses that affect humans (HCoV) can produce clinical symptoms from the common cold to serious ones like those caused by the Severe Acute Respiratory Syndrome (SARS) viruses and Middle East Respiratory Syndrome (MERS–CoV) [3]. The transmission mechanisms of SARS-COV-2 are animal–human and human–human. The first one is still unknown, but some researchers affirm that it could be through respiratory secretions and/or material from the digestive system. The second one, is considered similar for other Coronaviruses through the secretions of infected people, mainly by direct contact with respiratory drops and hands or fomites contaminated with these secretions, followed by contact with the mucosa of the mouth, nose or eyes [4].

Modeling is a science of creative capabilities connected with a profound learning in a variety of strategies to represent physical phenomena in the form of mathematical relations. In the prevailing situation, agencies, which control the diseases and maintain all the data of diseases, are publishing data of Covid-19 on daily bases. This data includes number of people having positive corona test, number of deaths, number of recoveries and active number of cases, and also commulative data from all over the world. So, appropriate model, with much accuracy, is needed at this level. Low dimensional models, with small number of compartments and having parameters which can be determined with the real data with good precision, are better to study and forecast the pandemic [5]. A high dimensions model requires a huge number of parameters to describe it but this huge number of parameters can not be found with enough precision [6]. In the absence of details, compartmental epidemic models describing the average behavior of the system can be a starting point. Even the simplest models contain several variables, which are hard to determine from the available data. The minimal *SIR* model describes the behavior of the susceptible *S*(*t*), the infected *I*(*t*), and the removed (recovered or deceased) *R*(*t*) populations [7,8]. Numerous models have been published on COVID-19 [9–15]. To the best of our knowledge, it has not been focused on the implications of mathematical model to guess the future trend of COVID-19 disease in Romania as well as in Pakistan. Thus the present study is taken to fill this gap.

To estimate the early dynamics of COVID-19 infection in Romania and Pakistan, we modeled the transmission through a deterministic *SIR* model. We are choosing the *SIR* model because in present situation worldwide data contains the infectious patients, recovered and deaths only, so from that data we can have average death rate and recovery. We estimate the size of the epidemic for both countries. We also forecast the maximum level of COVID-19 patients and time period for approaching the endemic level through model simulations. The dreadful effects of the pandemic, if precautionary measures or social distancing were ended, has also been analysed. We also perform the sensitivity analysis of the parameters by varying the values of transmission rate, disease-related death rate, recovery rate and the inhibition effect.

## 2 Structure of the Model

In an *SIR* type model, the total population is partitioned into three categories, the susceptible (S), the infectious (I), and the recovered (R). If the homogeneous mixing of people is assumed, the mathematical form of the model is given as:

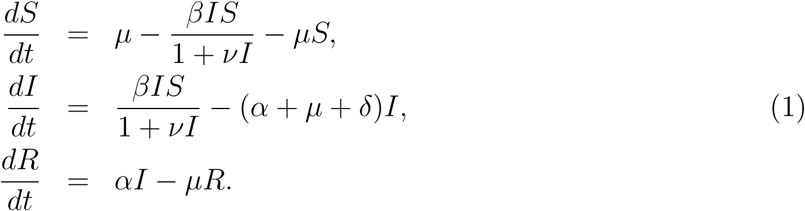

In the above model, we assume that birth and death rate is equal and is denoted by *μ*. The parameter *β* is the transmission rate as a result of the contact of susceptible individuals with the infected ones. The incidence term is assumed to be nonlinear and is represented as 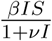. The parameter *ν* represents the inhibition effect or precautions that have been adopted to prevent the mixing of susceptible and infectious individuals. We assume that the recovery rate of infectious individuals is *α* and *δ* is the disease-related death rate.

## 3 Case Study for Romania

The coronavirus 2019-20 (COVID-19) pandemic was affirmed to have arrived in Romania on 26*^th^* February of this year [16]. Due to the spread of coronary disease in Italy, the government of Romania reported two weeks of isolation, starting from 21*^st^* February, for its residents which were coming back from the influenced regions [17]. On the very next day, the Romanian government declared a few preventive measures, including assignment of five clinics as separation-habitats for new cases, arrangement of warm scanners on airport terminals, and uniquely assigned lines for travelers originating from zones influenced by COVID-19 outbreak [18]. For avoiding the virus expansion several steps were taken by the government like on 9*^th^* March the authorities reported discontinuance of trips to and from Italy via all terminals [19] also the Special National Emergency Situations Committee ordered to close all schools on the same day. Two days later, on 11*^th^* March, the Government distributed a rundown of the fifteen rules in regards to the mindful social conduct in forestalling the spread of COVID-19 [20]. Specialists have forced a prohibition on all religious, scientific, sports, social or diversion occasions with more than 100 members for the next three weeks.

The number of affected people crossed the first hundred at the end of the second week of March. The first three deaths were announced in Romania on 22*^nd^* March. All three deceased were already suffering from different diseases such as diabetes, dialysis, and lung cancer, etc. [21]. Following a flood of new affirmed cases, on March 24, the administration declared Military Ordinance, establishing a national lock-down and bringing in the military to help police and the Gendarmerie in authorizing the new limitations. Developments outside the homes was strictly prohibited, with certain exemptions (work, purchasing nourishment or medication, and so forth.). Old people over 65 years were permitted to leave their homes just between 11 a.m.-1 p.m. [22]. Two days after this, on March 26, the National airline also suspended all local flights [23].

Total population of the Romania is about 19, 237, 691 [24]. The average life expectancy for people of Romania is 76 years [25]. One can see from the model (1) that we are involving disease-related death and immunity, so we have to fit our model with active real cases, active means no disease-related death, and no recovery. So, initially, we have 3 active cases on March 5, 2020. Hence, our initial conditions are *I*(0) = 3 and *R*(0) = 3 and rest are the susceptible. We have simulated our model and fit with the real cases. Figure (2) portrays the fitting of our model (1) with the real data given in figure (1).

**Figure 1:**
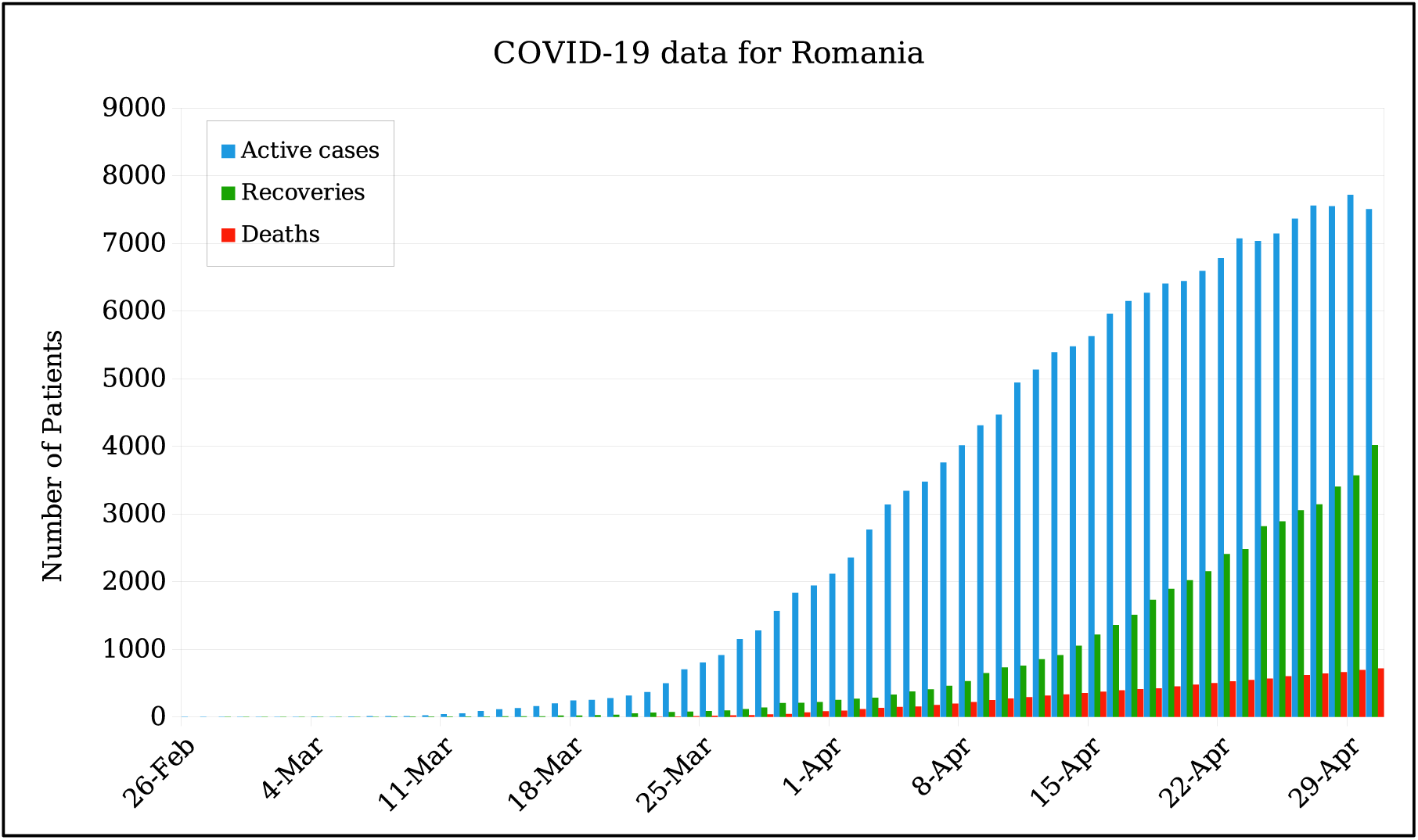
Real data of number of cumulative cases of COVID-19, per day, for Romania.

By observing the figure (2), one can compare the actual data reported by [26] and the data collected by the simple *SIR* model (1) given in section 2. We can see number of active cases are almost matching with the actual ones. We also estimate number of COVID-19 patients that will appear in next duration. It can be observed, from figure (2), that infection is continuously spreading un till August, 2020. After this period the malady is going to stable under the current situation. Note that here we have taken the average rate and disease related death rate per day up to April 30,2020. According to our estimate, there is no chance of vanishing the disease from community if the average daily and unfortunately disease related death rate is going on with the same rate. From figure (2), we can see that the number of patients will be ~ 10091 by the 31*^st^* May, on June 28*^th^* patients will be ~ 11127 and by the end of this year number will reach at ~ 12000. Week wise expected number of patients for the next months of this year is shown in Table 1.

**Figure 2:**
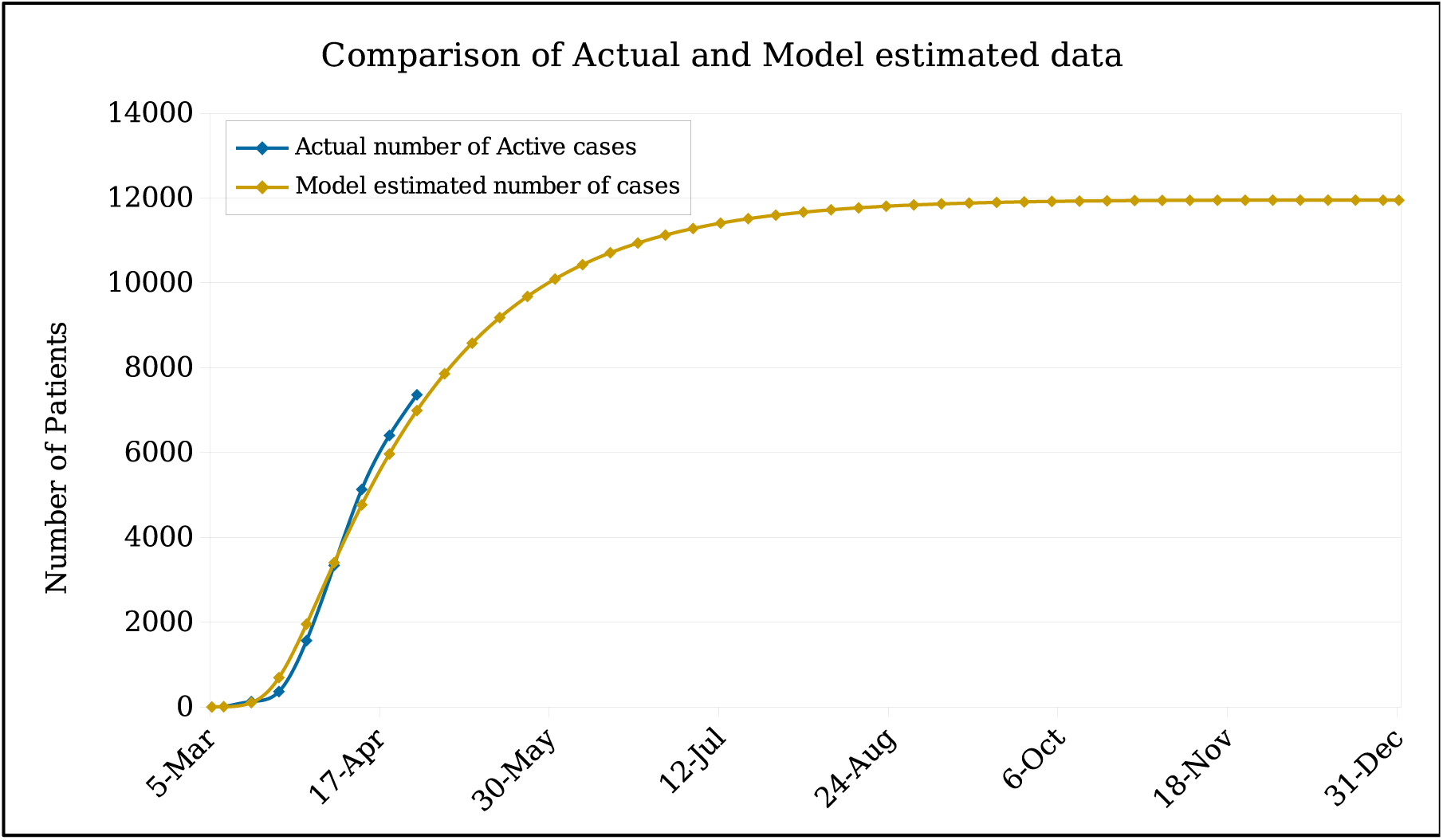
Comparison of the actual data of active COVID-19 patients with the model estimated number of patients and Forecasting the number of COVID-19 patients till December, 2020.

**Table 1:** Weekly Expected Number of active cases in Romania for the next months according to current situation

| Date | Estimated Number of Patients | Date | Estimated Number of Patients | Date | Estimated Number of Patients |
| --- | --- | --- | --- | --- | --- |
| 03-May | 7858 | 26-Jul | 11597 | 18-Oct | 11935 |
| 10-May | 8581 | 02-Aug | 11666 | 25-Oct | 11940 |
| 17-May | 9182 | 09-Aug | 11723 | 01-Nov | 11943 |
| 24-May | 9680 | 16-Aug | 11769 | 08-Nov | 11946 |
| 31-May | 10091 | 23-Aug | 11806 | 15-Nov | 11948 |
| 07-Jun | 10430 | 30-Aug | 11837 | 22-Nov | 11949 |
| 14-Jun | 10709 | 06-Sep | 11861 | 29-Nov | 11950 |
| 21-Jun | 10939 | 13-Sep | 11881 | 06-Dec | 11950 |
| 28-Jun | 11127 | 20-Sep | 11897 | 13-Dec | 11950 |
| 05-Jul | 11282 | 27-Sep | 11910 | 20-Dec | 11950 |
| 12-Jul | 11408 | 04-Oct | 11920 | 27-Dec | 11949 |
| 19-Jul | 11512 | 11-Oct | 11929 | 31-Dec | 11949 |

### 3.1 Variation in the Number of Patients with the Variation of Parameters

According to reported data, it has been observed that average weekly recovery rate and disease related death rate vary. The maximum average recovery rate happened between (1 – 7) March and it is 5.71%. During the week (29 March-4 April), minimum average recovery rate has been observed and its value is 3.5%. Similarly average disease related death rate varies every week. Its minimum value occurred between (12 April-18 April) which is 0.32%. The maximum average number of deaths per day appeared during the week (29 March-4 April) and its value is 0.7%. We vary the values of recovery and disease related death rates by observing this pattern and estimate the number of patients that will appear in the later weeks of this year. Similarly we increase and decrease the values of transmission rate and inhibition effect up to 25% and 50% and also estimate the number of COVID-19 cases. The effect of the transmission rate (*β*), the death rate due to COVID-19 (*δ*), recovery rate (*α*) and the inhibition or precautionary measures (*v*) on the number of COVID-19 patients have been calculated and shown in Figure 3.

**Figure 3:**
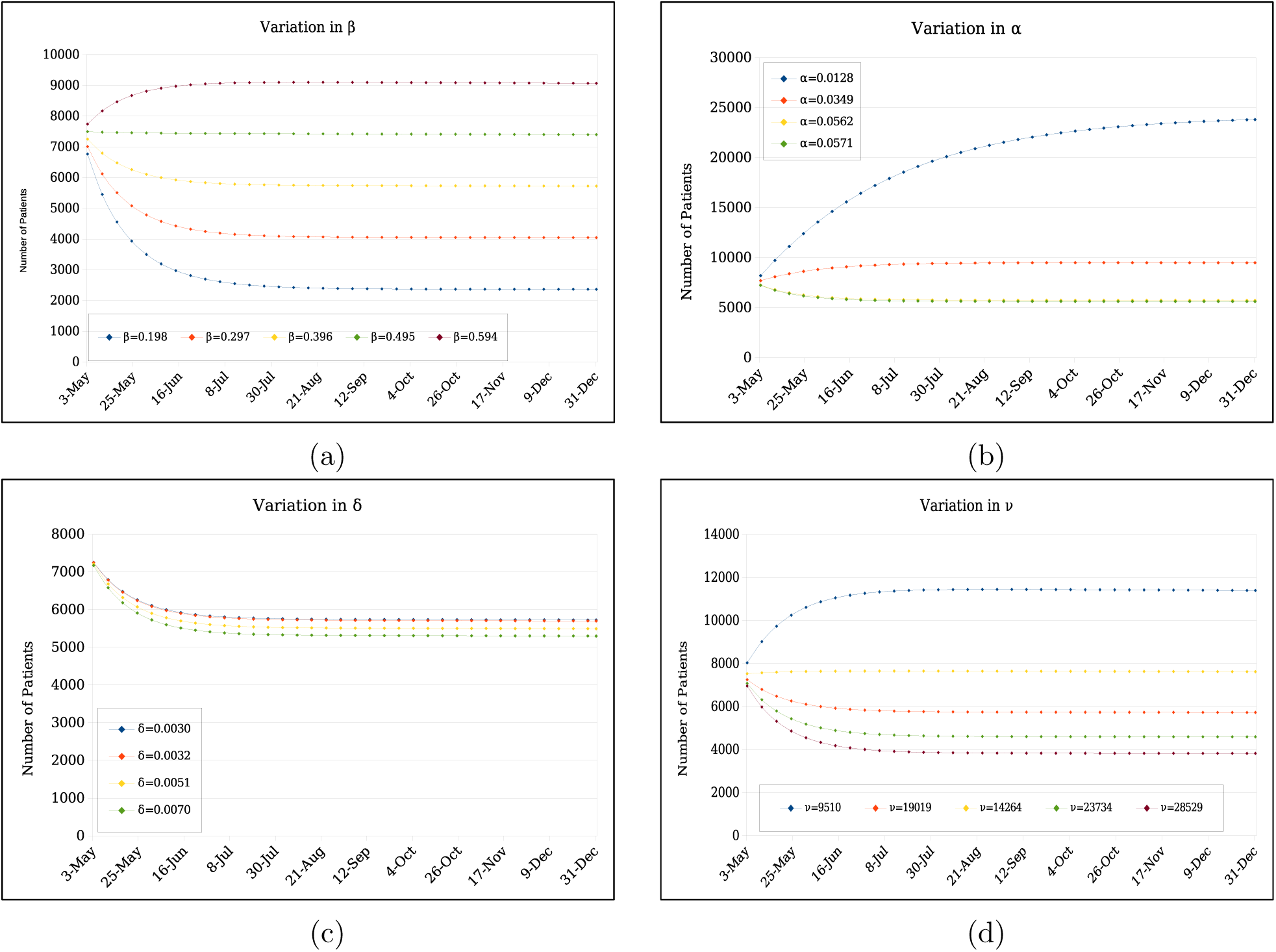
Variation in the number of active patients on transmission rate *β*, recovery rate *α*, death rate *δ* and the inhibition effect *ν*.

In Figure 3a, we present the dependency of the number of patients on the transmission rate *β*. The transmission rate is measured by the number of people get infected due to a source of COVID-19. For example *β* = 0.1 means every 10% people, per day, get infected. We can see from Figure 3a that the number of patients accelerates as *β* increases. The model fitted value for *β* is 0.396 and for that value, the number of patients by the end of this year will be ~ 12000. Since the transmission rate may vary for next duration, so we have estimated the number of patients by varying the value of *β* upto 25% and 50%. For *β* = 0.2, the number of patients by the end of this year decreases to ~ 2364. For *β* = 0.3, this number will be ~ 4046. For *β* = 0.5 the number of patients will be ~ 7400 and for *β* = 0.6 the number of patients will be ~ 9100. Week wise number of patients for each value of *β* are given in Table 2.

**Table 2:** Weekly Expected Number of patients, for Romania, for the next months for different values of *β*

| Date | $\beta = 0.198$ | $\beta = 0.297$ | $\beta = 0.396$ | $\beta = 0.495$ | $\beta = 0.594$ |
| --- | --- | --- | --- | --- | --- |
| 03-May | 6766 | 7009 | 7252 | 7497 | 7742 |
| 10-May | 5452 | 6120 | 6796 | 7479 | 8169 |
| 17-May | 4554 | 5507 | 6480 | 7467 | 8465 |
| 24-May | 3934 | 5082 | 6260 | 7458 | 8670 |
| 31-May | 3499 | 4784 | 6106 | 7451 | 8811 |
| 07-Jun | 3191 | 4574 | 5999 | 7446 | 8907 |
| 14-Jun | 2971 | 4426 | 5923 | 7442 | 8974 |
| 21-Jun | 2811 | 4321 | 5870 | 7439 | 9018 |
| 28-Jun | 2696 | 4245 | 5832 | 7436 | 9049 |
| 05-Jul | 2611 | 4192 | 5806 | 7434 | 9069 |
| 12-Jul | 2548 | 4153 | 5787 | 7432 | 9082 |
| 19-Jul | 2502 | 4125 | 5773 | 7431 | 9091 |
| 26-Jul | 2468 | 4105 | 5764 | 7429 | 9096 |
| 02-Aug | 2442 | 4091 | 5757 | 7427 | 9099 |
| 09-Aug | 2423 | 4081 | 5751 | 7426 | 9101 |
| 16-Aug | 2409 | 4073 | 5747 | 7424 | 9101 |
| 23-Aug | 2398 | 4067 | 5744 | 7423 | 9101 |
| 30-Aug | 2390 | 4063 | 5742 | 7422 | 9100 |
| 06-Sep | 2384 | 4060 | 5740 | 7420 | 9099 |
| 13-Sep | 2380 | 4058 | 5739 | 7419 | 9098 |
| 20-Sep | 2376 | 4056 | 5737 | 7417 | 9096 |
| 27-Sep | 2374 | 4054 | 5736 | 7416 | 9094 |
| 04-Oct | 2372 | 4053 | 5735 | 7415 | 9092 |
| 11-Oct | 2370 | 4052 | 5734 | 7413 | 9090 |
| 18-Oct | 2369 | 4051 | 5733 | 7412 | 9089 |
| 25-Oct | 2368 | 4051 | 5732 | 7411 | 9087 |
| 01-Nov | 2367 | 4050 | 5731 | 7409 | 9085 |
| 08-Nov | 2367 | 4050 | 5730 | 7408 | 9083 |
| 15-Nov | 2366 | 4049 | 5729 | 7407 | 9081 |
| 22-Nov | 2366 | 4049 | 5729 | 7405 | 9079 |
| 29-Nov | 2365 | 4048 | 5728 | 7404 | 9077 |
| 06-Dec | 2365 | 4048 | 5727 | 7403 | 9075 |
| 13-Dec | 2365 | 4047 | 5726 | 7401 | 9073 |
| 20-Dec | 2365 | 4047 | 5725 | 7400 | 9071 |
| 27-Dec | 2364 | 4046 | 5724 | 7399 | 9069 |
| 31-Dec | 2364 | 4046 | 5724 | 7398 | 9068 |

We next present our results, in Figure 3c, for the death rate dependence (*δ*) of the total number of COVID-19 patients. *δ* it is the total number of patients who died, per day, due to COVID-19 disease. *δ* = 0.001 means one patient dies, per day, in every thousand patients. Since all the other parameters are fixed, the trend of *δ* dependence is: higher the *δ* lower the number of active patients. As we know that *δ* varies day by day so we have plotted for five different values of *δ* ranging from 0.003 to 0.006 as the model fitted value of *δ* turns out to be 0.003. The total number of active patients by the end of this year ranges from 7000 – 6000 for this range of *δ*. Week wise number of active patients for the different values of *δ* are given in Table 3.

**Table 3:** Weekly Expected Number of patients, for Romaina, for the next months for different values of *α*

| Date | $\alpha = 0.0128$ | $\alpha = 0.0349$ | $\alpha = 0.0562$ | $\alpha = 0.0571$ |
| --- | --- | --- | --- | --- |
| 03-May | 8198 | 7699 | 7252 | 7232 |
| 10-May | 9723 | 8083 | 6796 | 6742 |
| 17-May | 11123 | 8386 | 6480 | 6404 |
| 24-May | 12401 | 8626 | 6260 | 6171 |
| 31-May | 13562 | 8816 | 6106 | 6009 |
| 07-Jun | 14616 | 8965 | 5999 | 5896 |
| 14-Jun | 15571 | 9082 | 5923 | 5817 |
| 21-Jun | 16433 | 9175 | 5870 | 5762 |
| 28-Jun | 17213 | 9247 | 5832 | 5723 |
| 05-Jul | 17916 | 9304 | 5806 | 5696 |
| 12-Jul | 18551 | 9348 | 5787 | 5676 |
| 19-Jul | 19123 | 9383 | 5773 | 5663 |
| 26-Jul | 19638 | 9410 | 5764 | 5653 |
| 02-Aug | 20103 | 9431 | 5757 | 5646 |
| 09-Aug | 20521 | 9447 | 5751 | 5640 |
| 16-Aug | 20896 | 9459 | 5747 | 5636 |
| 23-Aug | 21235 | 9469 | 5744 | 5633 |
| 30-Aug | 21539 | 9476 | 5742 | 5631 |
| 06-Sep | 21814 | 9481 | 5740 | 5629 |
| 13-Sep | 22058 | 9485 | 5739 | 5628 |
| 20-Sep | 22279 | 9488 | 5737 | 5626 |
| 27-Sep | 22477 | 9490 | 5736 | 5625 |
| 04-Oct | 22656 | 9491 | 5735 | 5624 |
| 11-Oct | 22816 | 9491 | 5734 | 5623 |
| 18-Oct | 22960 | 9492 | 5733 | 5622 |
| 25-Oct | 23089 | 9491 | 5732 | 5621 |
| 01-Nov | 23205 | 9491 | 5731 | 5620 |
| 08-Nov | 23308 | 9491 | 5730 | 5620 |
| 15-Nov | 23403 | 9490 | 5729 | 5619 |
| 22-Nov | 23485 | 9489 | 5729 | 5618 |
| 29-Nov | 23560 | 9488 | 5728 | 5617 |
| 06-Dec | 23626 | 9487 | 5727 | 5616 |
| 13-Dec | 23685 | 9486 | 5726 | 5615 |
| 20-Dec | 23739 | 9484 | 5725 | 5615 |
| 27-Dec | 23787 | 9483 | 5724 | 5614 |
| 31-Dec | 23812 | 9482 | 5724 | 5613 |

In Figure 3b, we present our results for the change in the total number of active patients as a function of recovery rate of infected patients *α*. As for the *β* and *δ*, *α* is also measured as a ratio per day. *α* = 0.01 means everyone out of hundred COVID-19 patients get recovered, per day. Definition of *α* infers the trend of the number of patients as a function of α: higher the value of *α* means lower the number of active COVID-19 patients. The model fitted value of *α* is 0.056. In Figure 3b, we have plotted for five different values of *α* including the model fitted one also. The other values of *α* that we have chosen are *α* = 0.013, 0.056, 0.058. The total number of active patients by the end of this year ranges from ~ 5613 to ~ 23812. Weekly details of the number of patients as a function of *α* is given in Table 4.

**Table 4:** Weekly Expected Number of patients, for Romania, for the next months for different values of *δ*

| Date | $\delta = 0.0030$ | $\delta = 0.0032$ | $\delta = 0.0051$ | $\delta = 0.0070$ |
| --- | --- | --- | --- | --- |
| 03-May | 7252 | 7248 | 7210 | 7171 |
| 10-May | 6796 | 6785 | 6682 | 6581 |
| 17-May | 6480 | 6464 | 6321 | 6182 |
| 24-May | 6260 | 6241 | 6074 | 5912 |
| 31-May | 6106 | 6086 | 5903 | 5727 |
| 07-Jun | 5999 | 5977 | 5784 | 5600 |
| 14-Jun | 5923 | 5901 | 5702 | 5513 |
| 21-Jun | 5870 | 5847 | 5645 | 5453 |
| 28-Jun | 5832 | 5810 | 5605 | 5411 |
| 05-Jul | 5806 | 5783 | 5577 | 5383 |
| 12-Jul | 5787 | 5764 | 5557 | 5363 |
| 19-Jul | 5773 | 5750 | 5544 | 5348 |
| 26-Jul | 5764 | 5741 | 5534 | 5338 |
| 02-Aug | 5757 | 5733 | 5526 | 5331 |
| 09-Aug | 5751 | 5728 | 5521 | 5326 |
| 16-Aug | 5747 | 5724 | 5517 | 5322 |
| 23-Aug | 5744 | 5721 | 5514 | 5320 |
| 30-Aug | 5742 | 5719 | 5512 | 5317 |
| 06-Sep | 5740 | 5717 | 5510 | 5316 |
| 13-Sep | 5739 | 5716 | 5509 | 5314 |
| 20-Sep | 5737 | 5714 | 5507 | 5313 |
| 27-Sep | 5736 | 5713 | 5506 | 5312 |
| 04-Oct | 5735 | 5712 | 5505 | 5311 |
| 11-Oct | 5734 | 5711 | 5504 | 5310 |
| 18-Oct | 5733 | 5710 | 5503 | 5309 |
| 25-Oct | 5732 | 5709 | 5502 | 5308 |
| 01-Nov | 5731 | 5708 | 5502 | 5307 |
| 08-Nov | 5730 | 5707 | 5501 | 5307 |
| 15-Nov | 5729 | 5706 | 5500 | 5306 |
| 22-Nov | 5729 | 5706 | 5499 | 5305 |
| 29-Nov | 5728 | 5705 | 5498 | 5304 |
| 06-Dec | 5727 | 5704 | 5498 | 5303 |
| 13-Dec | 5726 | 5703 | 5497 | 5303 |
| 20-Dec | 5725 | 5702 | 5496 | 5302 |
| 27-Dec | 5724 | 5701 | 5495 | 5301 |
| 31-Dec | 5724 | 5701 | 5495 | 5301 |

In Figure 3d, we present our results for the number of patients as a function of the inhibitory effect *ν*. The model fitted value of *ν* is 19019.1. Since this number can also vary, we have taken four other values of *ν* in Figure 3d. Since *ν* is proportional to the precautionary measures adopted by the COVID-19 patients along with the general population, higher values of *ν* means lower the number of active patients. The values that we have chosen for *ν* other than the model fitted value are *ν* = 9509.6, 14264.3, 23733.9. We can see in Figure 3d that the total number of COVID-19 patients ranges from 4591 – 11395. Weekly data for the number of COVID-19 patients as a function of five different values of *ν* is given in Table 5.

**Table 5:**
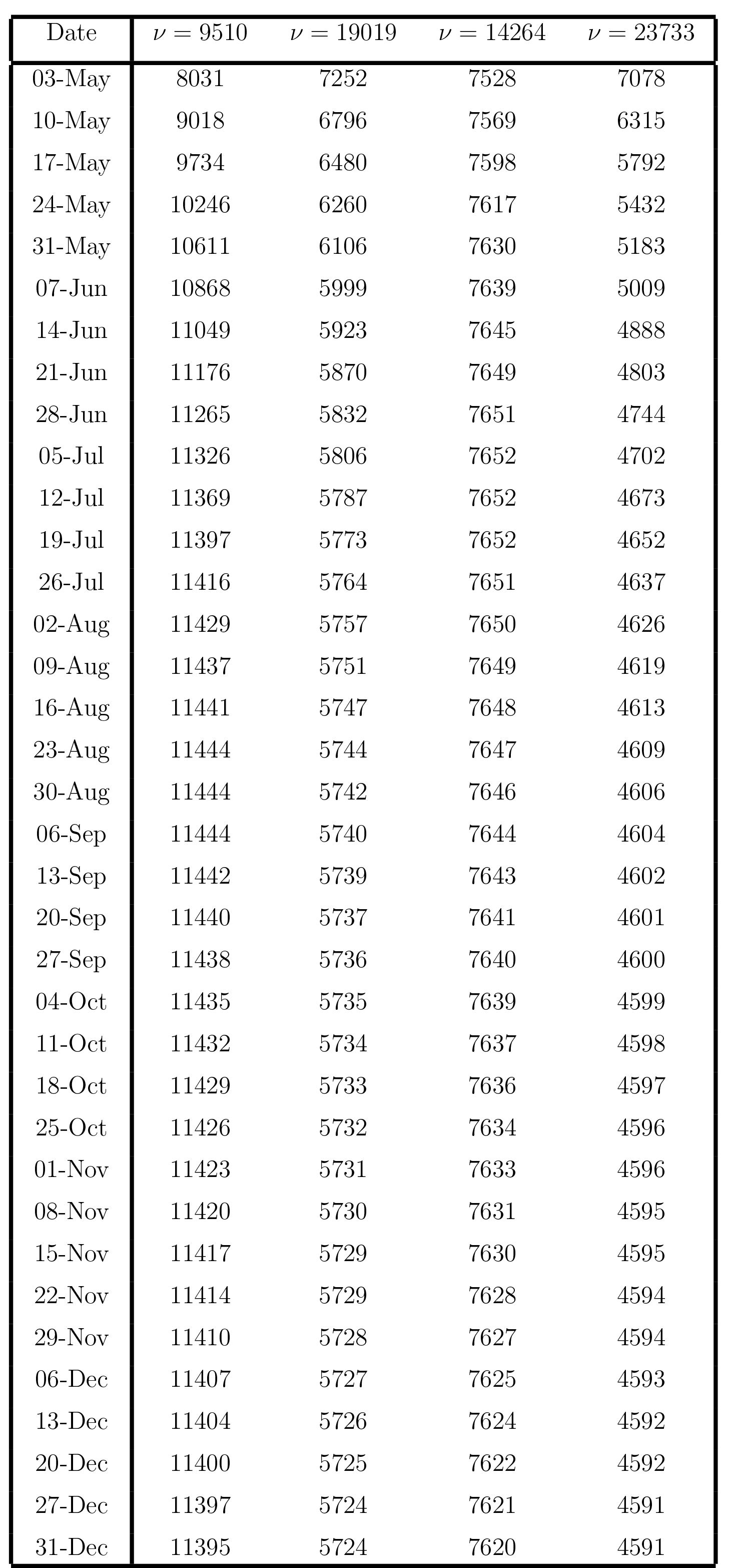
Weekly Expected Number of patients, for Romania, for the next months for different values of *ν*

**Table 6:** Weekly Expected Number of patients for the next months, in Romania, with the removal of all barriers

| Date | $\nu = 0$ | Date | $\nu = 0$ | Date | $\nu = 0$ |
| --- | --- | --- | --- | --- | --- |
| 03-May | 20592 | 26-Jul | 494332 | 18-Oct | 3857 |
| 10-May | 214423 | 02-Aug | 328907 | 25-Oct | 2585 |
| 17-May | 1971863 | 09-Aug | 218925 | 01-Nov | 1734 |
| 24-May | 8567506 | 16-Aug | 145779 | 08-Nov | 1163 |
| 31-May | 10657488 | 23-Aug | 97125 | 15-Nov | 781 |
| 07-Jun | 8115420 | 30-Aug | 64746 | 22-Nov | 525 |
| 14-Jun | 5596437 | 06-Sep | 43189 | 29-Nov | 353 |
| 21-Jun | 3768856 | 13-Sep | 28828 | 06-Dec | 238 |
| 28-Jun | 2518791 | 20-Sep | 19253 | 13-Dec | 160 |
| 05-Jul | 1678412 | 27-Sep | 12868 | 20-Dec | 108 |
| 12-Jul | 1117056 | 04-Oct | 8606 | 27-Dec | 73 |
| 19-Jul | 743114 | 11-Oct | 5760 | 31-Dec | 58 |

### 3.2 Dreadful Effects of Removal of Social Distancing and Precautionary Measures

According to the present recovery rate, disease-related death rate, and estimated values of transmission rate, we observe that if we remove the social distancing and adopted precautionary measures then the worst effects appear in the population. Almost ~ 55% population will be infected up to 31*^st^* May and then infected people will begin to decrease. Note that this situation will according to the current position. It means that it will happen only according to current transmission rate, recovery rate, disease related death rate. However, the situation may vary with the variation of these parameters. The epidemic curve without any barrier is shown in Fig. 4.

**Figure 4:**
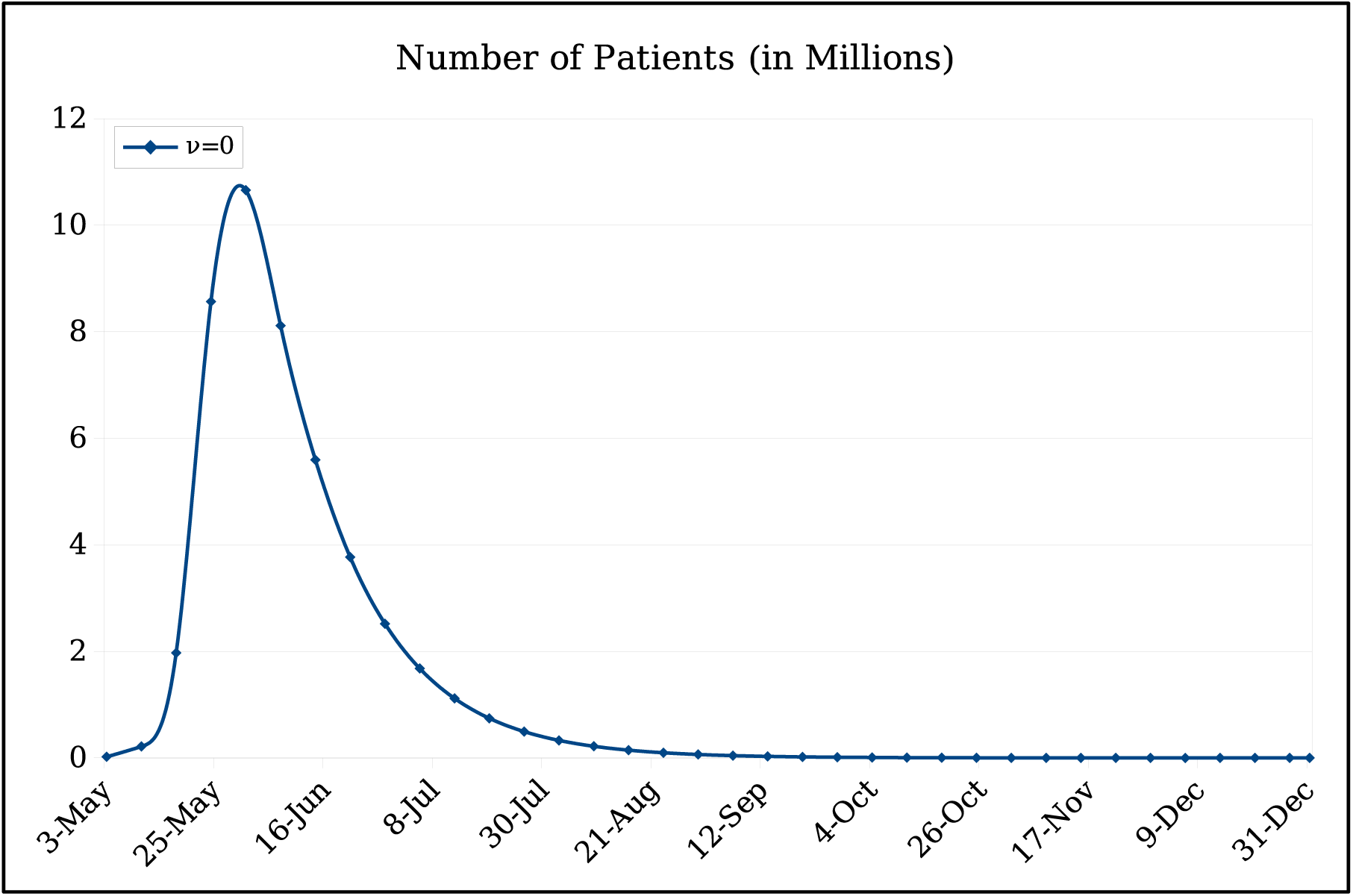
Epidemic curve of COVID-19 patients in Romania.

## 4 COVID-19 Case Study in Pakistan

The novel Coronavirus (COVID-19) pandemic was affirmed to have arrived at Pakistan on February 26, 2020. The first patient has been observed in Sindh Province and the second is in the federal territory of the country [27]. Within a week of appearance of initial two cases, this pandemic started to increase other areas of the country. On 29th April 2020, the quantity of affirmed cases in the nation is 15759, with 4052 (25.7% of the commulative cases)recuperation and 346 (2.2% of the commulative cases) deceased and Punjab is, right now, the area with the most elevated number of cases at over 6000 [28].

In figure (5), we have plotted only active cases with recovered and deaths from 26 of Feb, 2020 to 29 of April, 2020.

**Figure 5:**
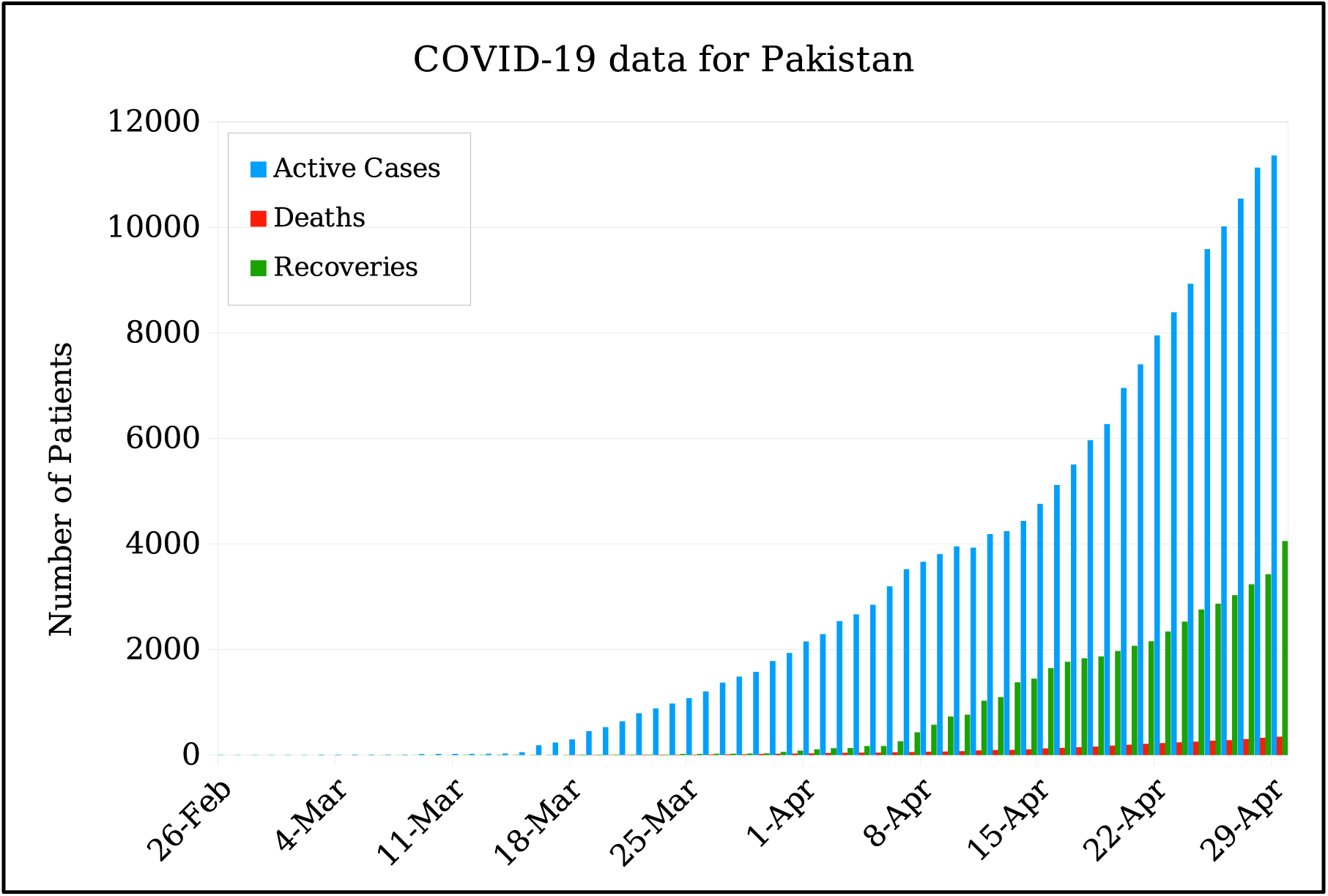
Real data of number of cumulative cases of COVID-19, per day, for Pakistan.

Currently Pakistan has, approximately, a total population of 220 millions [29] and life expectancy is 67 years [30]. As, we have included the disease related death and immunity in our proposed model (1) so this is telling us that we have to fit our model with the active cases of real data (deaths and recoveries are excluded), figure (6) is portraying the fitting of our model with real data, given in figure (5), from 1*^st^* of March, 2020 to 29 of April, 2020. The initial values are *I*(0) = 4, *R*(0) = 0 and the rest of the population is susceptible. In figure, we have compared week wise data and then extended this week wise data till 31 Dec., 2020 to forecast the Covid-19 cases in Pakistan. According to the figure (6), there will be ~ 30000 by the end of May, 2020 and at the end of August this number would be ~ 50000. Week wise expected number of patients for the next months of this year is shown in Table 7.

**Figure 6:**
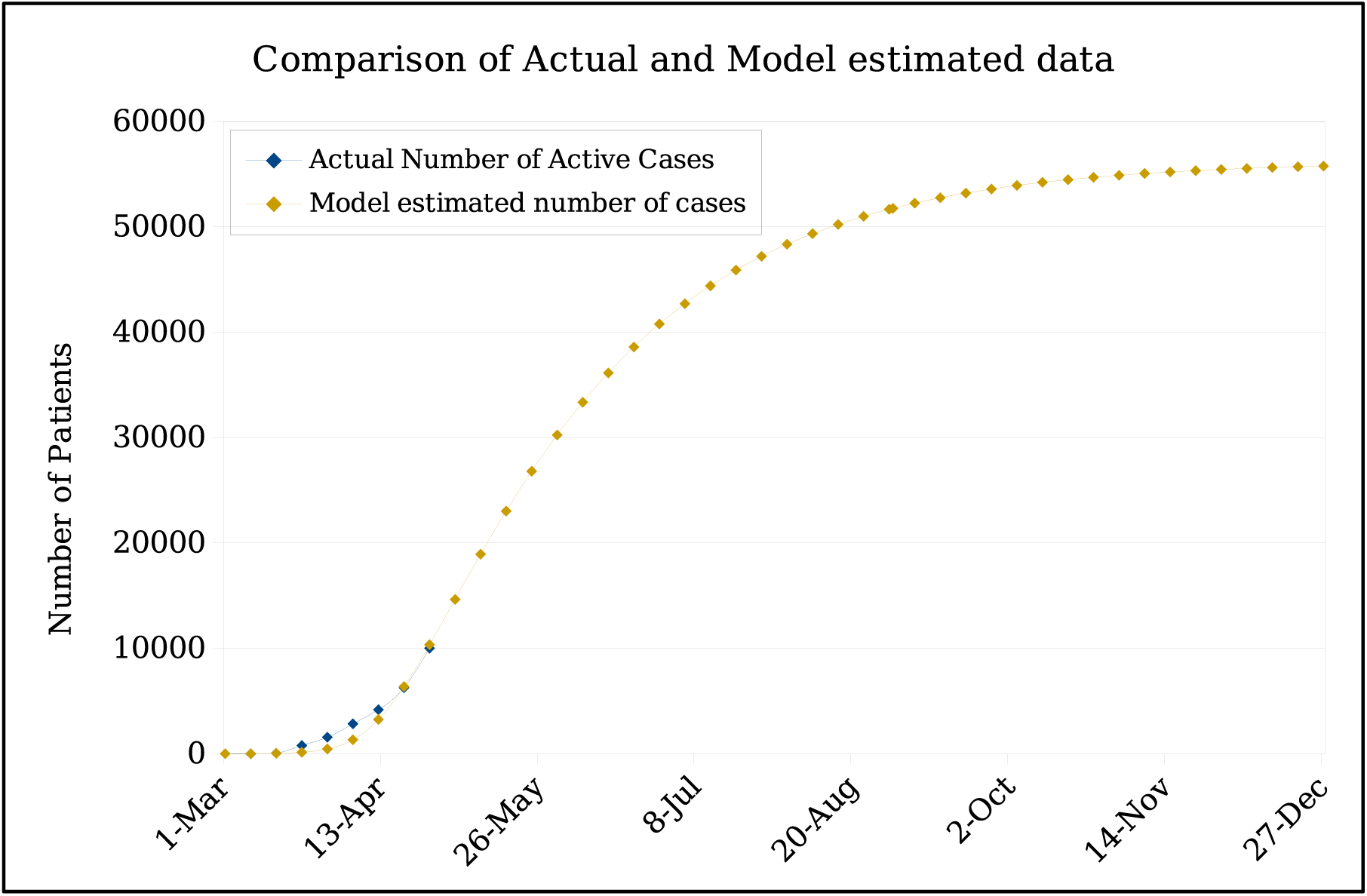
Comparison of actual data with estimated data and future prediction

**Table 7:** Weekly Expected Number of active cases, for Pakistan, for the next months according to current situation

| Date | Estimated Number of Cases | Date | Estimated Number of Cases |
| --- | --- | --- | --- |
| 03-May | 14652 | 06-Sep | 52257 |
| 10-May | 18944 | 13-Sep | 52767 |
| 17-May | 23030 | 20-Sep | 53211 |
| 24-May | 26814 | 27-Sep | 53599 |
| 31-May | 30261 | 04-Oct | 53937 |
| 07-Jun | 33365 | 11-Oct | 54230 |
| 14-Jun | 36142 | 18-Oct | 54487 |
| 21-Jun | 38608 | 25-Oct | 54710 |
| 28-Jun | 40790 | 01-Nov | 54903 |
| 05-Jul | 42717 | 08-Nov | 55073 |
| 12-Jul | 44414 | 15-Nov | 55218 |
| 19-Jul | 45905 | 22-Nov | 55345 |
| 26-Jul | 47212 | 29-Nov | 55458 |
| 02-Aug | 48358 | 06-Dec | 55552 |
| 09-Aug | 49361 | 13-Dec | 55636 |
| 16-Aug | 50237 | 20-Dec | 55708 |
| 23-Aug | 51005 | 27-Dec | 55770 |
| 30-Aug | 51674 | 31-Dec | 55803 |

### 4.1 Variation in the number of Covid-19 patients by changing the values of parameters

In this section, we will see that how the number of active cases of Covid-19 vary if we change the values of parameters. Figure (7) is depicting the effect of variations in parameters on the number of active Covid-19 cases.

**Figure 7:**
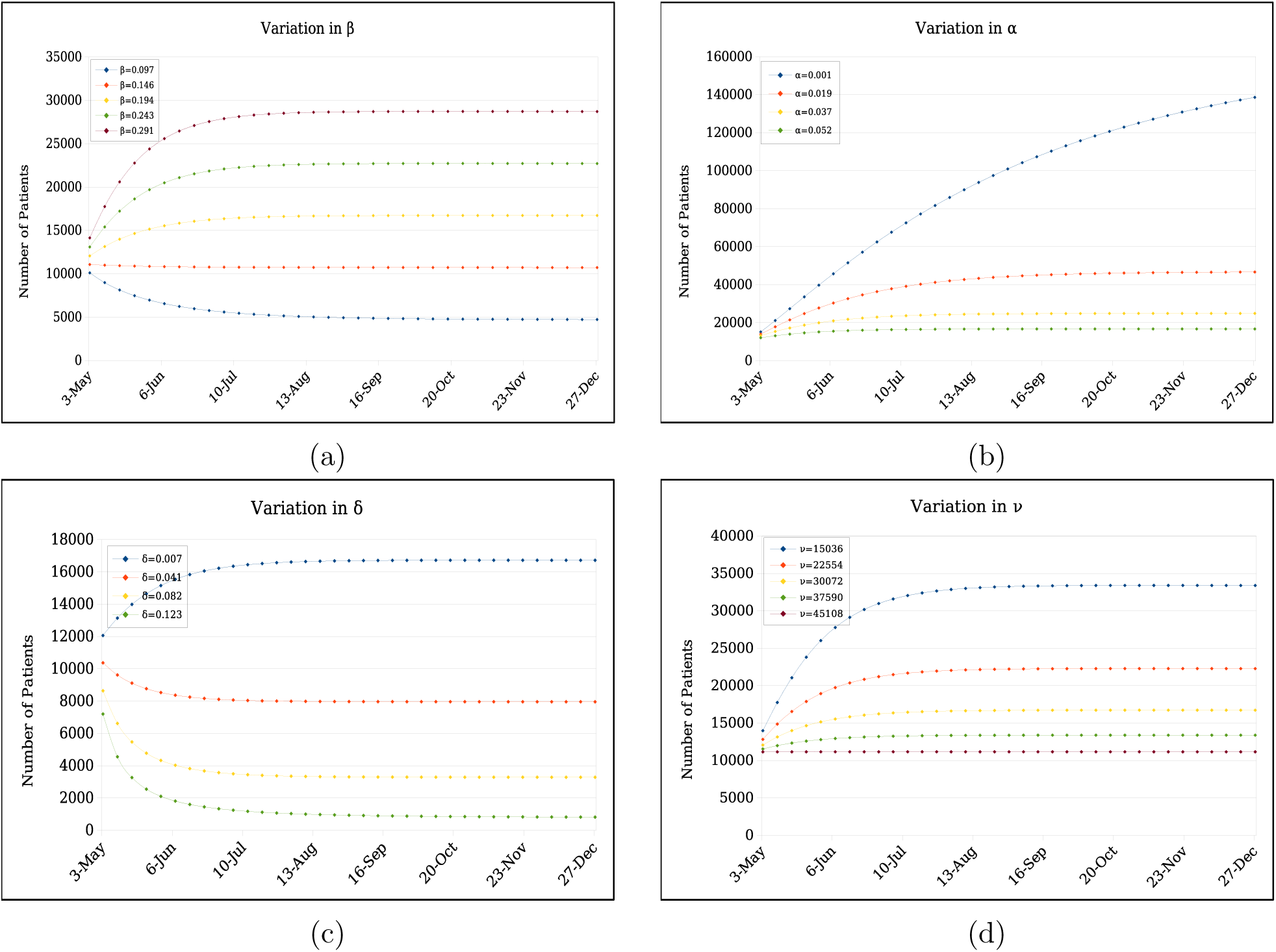
Variation in the number of active patients on transmission rate *β*, death rate *δ*, recovery rate *α* and the inhibition effect *ν*.

**Figure 8:**
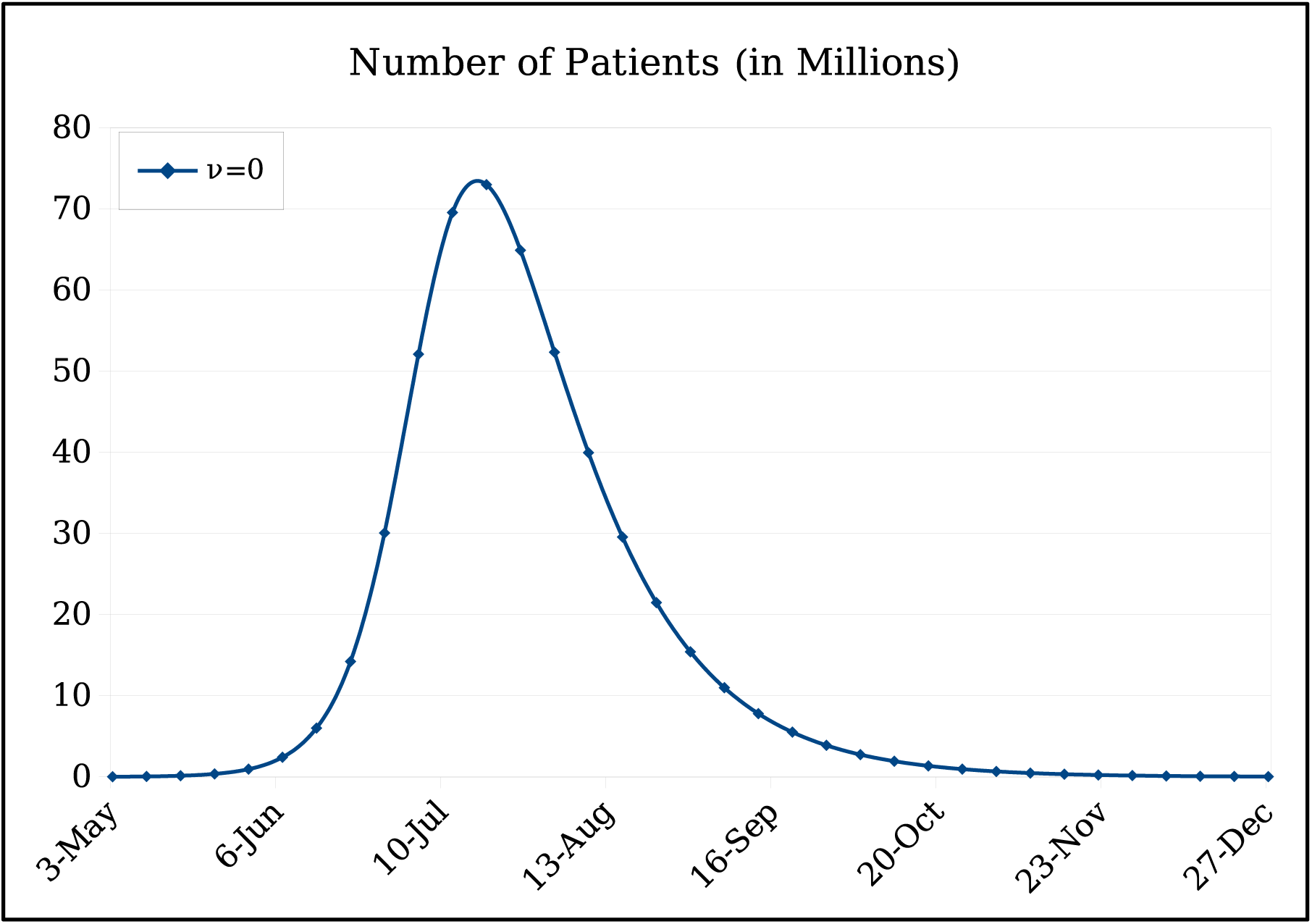
Epidemic curve of COVID-19 patients in Pakistan.

Figure 7a represents the dependence of number of patients on the variation of transmission rate *β*. This rate tells that how many people are getting infection per day. For example if *β* = 0.097 then it means that 97 people are getting infection per day per 1000 people. We have taken five different values of *β* including the model fitted value *β* = 0.194, we can see that by increasing the transmission rate number of cases are also increasing as expected, Table 8 contains all the possible number of patients for different values of *β*.

**Table 8:**
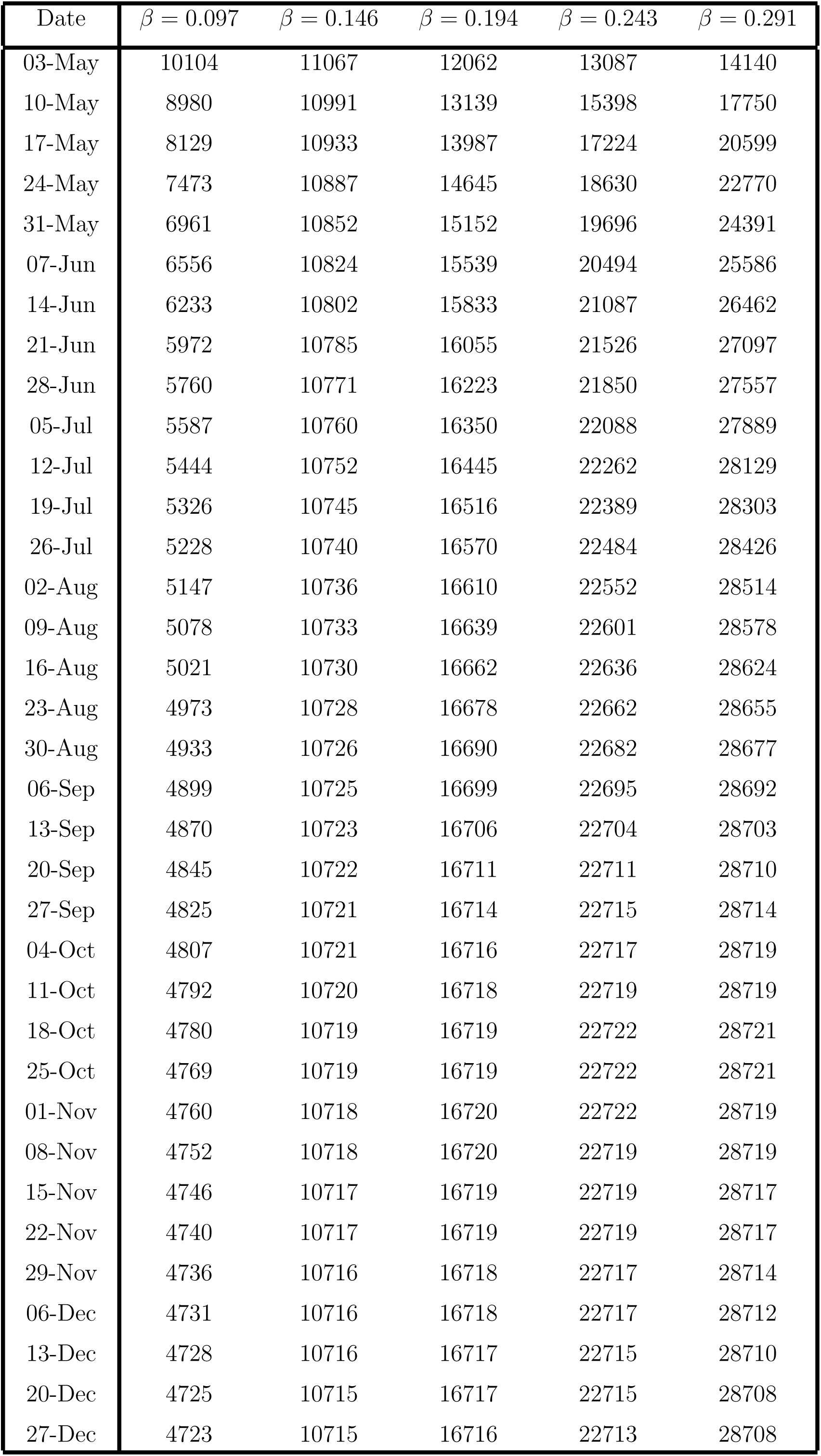
Weekly Expected Number of patients, for Pakistan, for the next months for dierent values of *β*

Next, we will check the dependence of number of active cases on recovery rate, *α*. It is the rate which tells that how many people are getting immunity from this disease. For example if *α* = 0.001 then it means that out of 1000 people one person is recoverd per day. We have taken four different values of *α*, one is our model fitted value which is *α* = 0.015 and three from the real data [28], by observing the real data we perceived that average recovery rate is maximum for the week 19*^th^* – 25*^th^* April, 2020 which is 0.037 and minimum for the week 15*^th^* – 21*^st^* April, 2020 which is 0.001, so we have considered these two values and fourth is the average of 0.037 and 0.001. Figure 7b represents the trend of active cases depending on *α*, we can see that number of Covid-19 cases is inversely proportional to the recovery rate *α*, which makes sense. All the possible number of cases for all these values of *α* are given in Table 9.

**Table 9:** Week wise data for the number of COVID-19 patients, for Pakistan, for four different values of *α*.

| Date | $\alpha = 0.001$ | $\alpha = 0.019$ | $\alpha = 0.037$ | $\alpha = 0.052$ |
| --- | --- | --- | --- | --- |
| 03-May | 15137 | 13972 | 13233 | 12062 |
| 10-May | 21150 | 17858 | 15393 | 13139 |
| 17-May | 27366 | 21493 | 17222 | 13987 |
| 24-May | 33603 | 24798 | 18740 | 14645 |
| 31-May | 39750 | 27753 | 19981 | 15152 |
| 07-Jun | 45742 | 30364 | 20986 | 15539 |
| 14-Jun | 51544 | 32652 | 21795 | 15833 |
| 21-Jun | 57134 | 34643 | 22442 | 16055 |
| 28-Jun | 62502 | 36373 | 22959 | 16223 |
| 05-Jul | 67639 | 37869 | 23368 | 16350 |
| 12-Jul | 72549 | 39158 | 23694 | 16445 |
| 19-Jul | 77233 | 40269 | 23951 | 16516 |
| 26-Jul | 81699 | 41224 | 24156 | 16570 |
| 02-Aug | 85947 | 42042 | 24317 | 16610 |
| 09-Aug | 89989 | 42744 | 24444 | 16639 |
| 16-Aug | 93832 | 43344 | 24543 | 16662 |
| 23-Aug | 97482 | 43859 | 24622 | 16678 |
| 30-Aug | 100947 | 44299 | 24684 | 16690 |
| 06-Sep | 104236 | 44675 | 24732 | 16699 |
| 13-Sep | 107356 | 44997 | 24770 | 16706 |
| 20-Sep | 110317 | 45272 | 24801 | 16711 |
| 27-Sep | 113122 | 45507 | 24823 | 16714 |
| 04-Oct | 115782 | 45707 | 24840 | 16716 |
| 11-Oct | 118303 | 45877 | 24856 | 16718 |
| 18-Oct | 120692 | 46022 | 24867 | 16719 |
| 25-Oct | 122956 | 46145 | 24873 | 16719 |
| 01-Nov | 125099 | 46251 | 24880 | 16720 |
| 08-Nov | 127129 | 46341 | 24884 | 16720 |
| 15-Nov | 129050 | 46418 | 24889 | 16719 |
| 22-Nov | 130871 | 46482 | 24891 | 16719 |
| 29-Nov | 132594 | 46537 | 24893 | 16718 |
| 06-Dec | 134224 | 46583 | 24893 | 16718 |
| 13-Dec | 135769 | 46622 | 24895 | 16717 |
| 20-Dec | 137229 | 46658 | 24895 | 16717 |
| 27-Dec | 138611 | 46684 | 24895 | 16716 |

Next, we will see that how the death rate *δ* effects the number of Covid-19 cases. It is the rate which tells that how many people die from this disease. For example if *δ* = 0.007 then it means that out of 1000 people seven people die per day. We have taken four different values of *δ*, one is our model fitted value which is *δ* = 0.00703844071 and three from the real data [28]. We have seen that average death rate is minimum for the week 19*^th^* – 25*^th^* April, 2020 which is 0.004 and maximum for the week 15*^th^* – 21*^st^* April, 2020 which is 0.00122985. Fourth is 0.0008, it is the average of 0.004 and 0.001. Figure 7c is depicting the number of active cases as a function of *δ*. In the Table 10, we have calculated the number of Covid-19 cases for all these values of *δ*. In Figure 7d, we present our results for the number of patients as a function of the inhibition effect *ν*. The model fitted value of *ν* is 30072. Since this number can also vary, we have taken four other values of *ν* in Figure 7d. Since *ν* is proportional to the precautionary measures adopted by the COVID-19 patients along with the general population, higher values of *ν* means lower the number of active patients. The values that we have chosen for *ν* other than the model fitted value are *ν* = 15036.1, 22554.2, 37590.2, 45108.3. We can see in Figure 7d that the total number of COVID-19 patients ranges from 5500 – 8000. The per day data for number of COVID-19 patients as a function of five different values of *ν* is given in Table 11.

**Table 10:** Week wise data for the number of COVID-19 patients, for Pakistan, for four different values of *α*.

| Date | $\delta = 0.007$ | $\delta = 0.040$ | $\delta = 0.082$ | $\delta = 0.123$ |
| --- | --- | --- | --- | --- |
| 03-May | 12062 | 10368 | 8638 | 7198 |
| 10-May | 13139 | 9617 | 6615 | 4551 |
| 17-May | 13987 | 9112 | 5469 | 3263 |
| 24-May | 14645 | 8767 | 4772 | 2545 |
| 31-May | 15152 | 8529 | 4324 | 2101 |
| 07-Jun | 15539 | 8362 | 4025 | 1807 |
| 14-Jun | 15833 | 8246 | 3819 | 1599 |
| 21-Jun | 16055 | 8164 | 3674 | 1448 |
| 28-Jun | 16223 | 8105 | 3570 | 1333 |
| 05-Jul | 16350 | 8064 | 3495 | 1244 |
| 12-Jul | 16445 | 8035 | 3441 | 1173 |
| 19-Jul | 16516 | 8014 | 3401 | 1117 |
| 26-Jul | 16570 | 7999 | 3371 | 1070 |
| 02-Aug | 16610 | 7988 | 3349 | 1032 |
| 09-Aug | 16639 | 7980 | 3333 | 1000 |
| 16-Aug | 16662 | 7975 | 3320 | 973 |
| 23-Aug | 16678 | 7970 | 3311 | 950 |
| 30-Aug | 16690 | 7967 | 3304 | 930 |
| 06-Sep | 16699 | 7965 | 3299 | 914 |
| 13-Sep | 16706 | 7963 | 3295 | 900 |
| 20-Sep | 16711 | 7962 | 3293 | 887 |
| 27-Sep | 16714 | 7961 | 3290 | 876 |
| 04-Oct | 16716 | 7960 | 3289 | 867 |
| 11-Oct | 16718 | 7959 | 3287 | 859 |
| 18-Oct | 16719 | 7959 | 3286 | 852 |
| 25-Oct | 16719 | 7958 | 3285 | 846 |
| 01-Nov | 16720 | 7958 | 3285 | 840 |
| 08-Nov | 16720 | 7957 | 3284 | 836 |
| 15-Nov | 16719 | 7957 | 3284 | 831 |
| 22-Nov | 16719 | 7957 | 3284 | 828 |
| 29-Nov | 16718 | 7956 | 3283 | 825 |
| 06-Dec | 16718 | 7956 | 3283 | 822 |
| 13-Dec | 16717 | 7955 | 3283 | 819 |
| 20-Dec | 16717 | 7955 | 3283 | 817 |
| 27-Dec | 16716 | 7955 | 3282 | 815 |

**Table 11:**
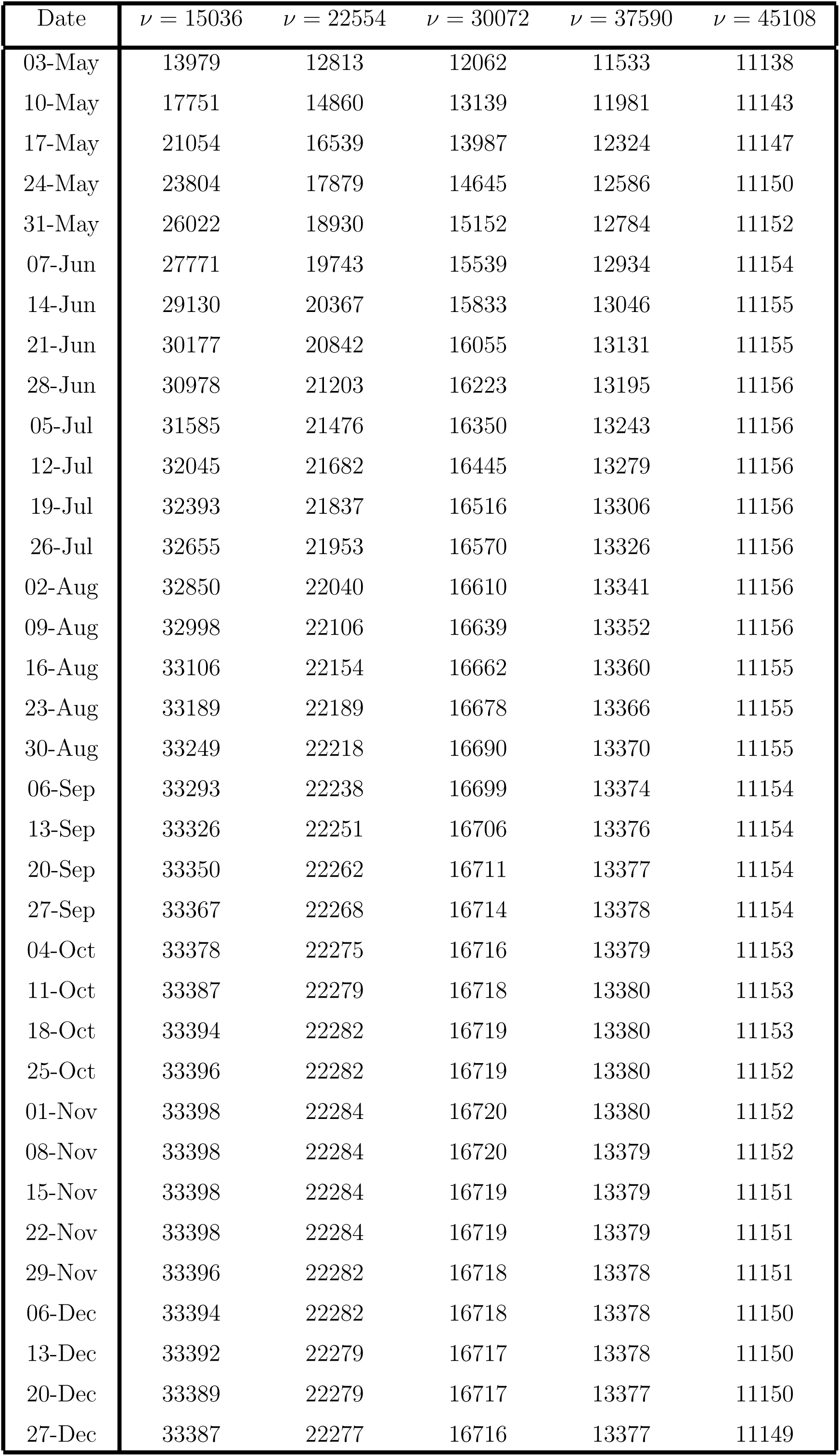
Week wise data for the number of COVID-19 patients, for Pakistan, for five different values of *ν*.

### 4.2 Dreadful Effects of Removal of Social Distancing and Precautionary Measures

We know that the major factor to aviod from the Covid-19 are social distancing and precautionary measures, in our model we have considered *ν* as this major factor. Now, if we have the present scenario and we consider do not take care of *ν* then we can see from the figure that almost 33% population of the whole country will be infected till 19*^th^* of July, 2020 and this is the peak of infection after this it will start decreasing, we have shown the calculated results are given in Table 12.

**Table 12:** Weekly Expected Number of patients for the next months, for Pakistan, with the removal of all barriers

| Date | $\nu = 0$ | Date | $\nu = 0$ | Date | $\nu = 0$ |
| --- | --- | --- | --- | --- | --- |
| 03-May | 21879 | 26-Jul | 64913200 | 18-Oct | 1364792 |
| 10-May | 56329 | 02-Aug | 52355600 | 25-Oct | 961312 |
| 17-May | 144949 | 09-Aug | 39971800 | 01-Nov | 677116 |
| 24-May | 372504 | 16-Aug | 29574600 | 08-Nov | 477004 |
| 31-May | 954074 | 23-Aug | 21479480 | 15-Nov | 336072 |
| 07-Jun | 2422860 | 30-Aug | 15422880 | 22-Nov | 236852 |
| 14-Jun | 6022720 | 06-Sep | 10994720 | 29-Nov | 166962 |
| 21-Jun | 14224540 | 13-Sep | 7801860 | 06-Dec | 117733 |
| 28-Jun | 30063000 | 20-Sep | 5519580 | 13-Dec | 83048 |
| 05-Jul | 52104800 | 27-Sep | 3897520 | 20-Dec | 58599 |
| 12-Jul | 69564000 | 04-Oct | 2748900 | 27-Dec | 41362 |
| 19-Jul | 73007000 | 11-Oct | 1937276 |  |  |

## 5 Conclusion

In this study, we used a mathematical model to assess the feasibility of the appearance of COVID-19 cases in Romania and Pakistan as well as the ultimate number of patients according to the current situation. By comparing model outcomes with the confirmed cases, it has been observed that our estimated values have good correspondence with the confirmed numbers. If the current pattern is going on then, according to our estimate, there will be ~ 12000 infectious individuals in Romania by the end of this year. Pakistan will bear the burden of ~ 55800 till the end of December, 2020. The situation will vary by the variation of transmission rate, death rate, recovery rate, and further implementation of social distancing in both countries. It has been observed that average weekly recovery rate and average weekly disease related death vary for both countries.

If the transmission rate in Romania increases 50% and recovery rate and disease related death rate is taken for 30*^th^* April, according to reported data, then there will be ~ 9000 persons carrying Corona malady and if this rate decreases 50%, then 2364 infected persons will exist in the Romanian community by the end of this year. If we take previous average maximum weekly recovery rate and disease related death rate, then there will be ~ 5613 and ~ 5301, patients, respectively in Romania. Similarly, by assuming minimum weekly average recovery and disease related death rate will result ~ 23812 and ~ 5724, respectively. The inhibition effect or precautionary measures also influence in the spreading of pandemic. If the inhibition factor increases upto 50%, then ~ 4951 patients will be existing in Romania till the end of this year. This number will exceed to ~ 11395, if precautionary measures decreases to 50%. The worst effects of the disease appear in the community, if we remove all the barriers. In such case, this malady may increase by effecting ~ 55% population till the end of this month. This number will start to decrease after May.

Increase or decrease in transmission rate will also results in decrease or increase in the number of COVID-19 patients in Pakistan. If the transmission rate increases 50% and recovery rate and disease related death rate is taken for 28*^th^* April, according to reported data, then there will be ~ 28708 persons having Corona disease and if this rate decreases 50%, then 4723 infected persons will exist among Pakistanis by the end of this year. If we take previous average maximum weekly recovery rate and disease related death rate, then there will be ~ 16716 and ~ 815, patients, respectively in Pakistan. Similarly, by assuming minimum weekly average recovery and disease related death rate will result ~ 138611 and ~ 16716, respectively. The inhibition effect or precautionary measures also influence in the spreading of pandemic. If the inhibition factor increases upto 50%, then ~ 11149 patients will be existing in Pakistan till the end of this year. This number will exceed to ~ 33387, if precautionary measures decreases to 50%. The worst effects of the disease appear in the community, if we remove all the barriers. In such case, this infection may increase by effecting ~ 33% population till the end of this month. This number will start to decrease after May, 2020.

Although these estimates may vary with the passage of time but it will really help us to observe the most influential factors that causes to increase the epidemic. On the basis of this analysis competent authorities may design the most effective strategies in order to control the epidemic.

## Data Availability

All real data used in the manuscript is publicaly available.

## References

[1] L. D. Izquierdo, et al., Informe tecnico nuevo coronavirus 2019–ncov, Ph.D. thesis, Instituto de Salud Carlos III (2020).

[2] W. H. Organization, Novel coronavirus (2019-ncov) situation reports. 2020. https://www.who.int/emergencies/diseases/novel-coronavirus-2019/situation-reports.

[3] C. I. Paules, H. D. Marston, A. S. Fauci, Coronavirus infections more than just the common cold, Jama 323 (8) (2020) 707–708.

[4] L. Saif, Animal coronavirus vaccines: lessons for sars., Developments in biologicals 119 (2004) 129–140. URL https://europepmc.org/article/med/15742624

[5] Jardón-Kojakhmetov H, Kuehn C, Pugliese A, Sensi M. A geometric analysis of the SIR, SIRS and SIRWS epidemiological models. arXiv preprint arXiv:2002.00354. 2020 Feb 2.

[6] J. T. Wu, K. Leung, G. M. Leung, Nowcasting and forecasting the potential domestic and international spread of the 2019-nCoV outbreak originating in Wuhan, China: a modelling study. The Lancet. 2020; 395(10225): 689–697. doi:10.1016/S0140-6736(20)30260-9.

[7] Kermack, W.O. and McKendrick, A.G., A contribution to the mathematical theory of epidemics. Proceedings of the royal society of London. Series A, 115(772), pp.700–721. (1927).

[8] Brauer F, Castillo-Chavez C, Feng Z. Mathematical Models in Epidemiology. Springer New York; 2019 Feb 20.

[9] A. J. Kucharski, et al. Early dynamics of transmission and control of COVID-19: a mathematical modelling study. Lancet Infect Dis. 2020; Online First.

[10] M. Kochan’czyk, F. Grabowski, and T. Lipniacki, Dynamics of COVID-19 pandemic at constant and time-dependent contact rates. Mathematical Modelling of Natural Phenomena, 15, 28, (2020). doi:10.1016/S1473-3099(20)30144-4.

[11] D. S. Hui et al. The continuing 2019-CoV epidemic threat of novel coronaviruses to global health-The latest 2019 novel coronavirus outbreak in Wuhan, China. Int J Infect Dis. 2020; 91: 264–266.

[12] M. Banerjee, A. Tokarev and V. Volpert, Immuno-epidemiological model of two-stage epidemic growth. Mathematical Modelling of Natural Phenomena, 15, 27,(2020).

[13] C. Anastassopoulou, L. Russo, A. Tsakris, C. Siettos, Data-based analysis, modelling and forecasting of the COVID-19 outbreak. MedRxiv. March 2020. doi:https://doi.org/10.1101/2020.02.11.20022186.

[14] K. Roosa et al. Real-time forecasts of the COVID-19 epidemic in China from February 5th to February 24th, 2020. Infect Dis Model. 2020; 5: 256–263. doi:10.1016/j.idm.2020.02.002.

[15] V. Volpert, M. Banerjee, and S. Petrovskii, On a quarantine model of coronavirus infection and data analysis. Mathematical Modelling of Natural Phenomena, 15, 24, (2020).

[16] Primul caz de coronavirus in Romania. Suspiciuni despre un al doilea caz in Gorj (First coronavirus case in Romania. Suspicions about a second case in Gorj). Digi24 (in Romanian). Retrieved 26th February 2020.

[17] Radio Romania International - Measures against the coronavirus. Radio Romania International. Archived from the original on 26th February 2020.

[18] Epidemia din China. Posibilitatea ca noul coronavirus sa ajunga in Romania (Epidemic from China. The possibility of the coronavirus to arrive in Romania). Stirile Pro TV (in Romanian). Archived from the original on 28th January 2020. Retrieved 27th January 2020.

[19] Romania suspenda zborurile din si catre Italia. www.digi24.ro (in Romanian). Retrieved 9th March 2020.

[20] 15 Recomandari privind conduita sociala responsabila in prevenirea raspandirii coronavirus (COVID-19). Ministry of Internal Affairs. 11th March 2020.

[21] Inca un romania murit din cauza conoravirusului. Trei decese confirmate in Romania (Another Romanian died because of coronavirus. Three deaths confirmed in Romania). Digi24 (in Romanian). Retrieved 22nd March 2020.

[22] Gogu, Madalina. “Military Ordinance No.3” (in Romanian). Retrieved 25th March 2020.

[23] Romanian state airline Tarom suspends all internal flights. Romania Insider. 26th March 2020. Retrieved 27th March 2020.

[24] https://www.macrotrends.net/countries/ROU/romania/population.

[25] https://www.macrotrends.net/countries/ROU/romania/life-expectancy.

[26] https://www.worldometers.info/coronavirus/country/romania.

[27] https://www.pakistantoday.com.pk/2020/02/26/sindh-health-two-coronavirus-cases-confirmed-in-pakistan-confirms-first-coronavirus-case-in-karachi.

[28] http://covid.gov.pk/

[29] https://www.macrotrends.net/countries/PAK/pakistan/population

[30] https://www.macrotrends.net/countries/PAK/pakistan/life-expectancy

